# The lifetime accumulation of multimorbidity and its influence on dementia risk: a UK Biobank Study

**DOI:** 10.1101/2024.01.21.24301584

**Authors:** R. Patel, C.E. Mackay, L. Griffanti, G. Gillis, K.P. Ebmeier, S. Suri

**Author notes:** **Corresponding Author:** Dr Raihaan Patel Oxford Centre for Human Brain Activity Wellcome Centre for Integrative Neuroimaging Department of Psychiatry University of Oxford Oxford, UK **Email:**.

## Abstract

The number of people living with dementia worldwide is projected to reach 150 million by 2050, making prevention a crucial priority for health services^1^. The co-occurrence of two or more chronic health conditions, termed multimorbidity, occurs in up to 80% of dementia patients^2^, raising the potential of multimorbidity as an important risk factor for dementia. However, precise understanding of which specific conditions, as well as their age of onset, drive the link between multimorbidity and dementia is unclear. We defined the patterns of accumulation of 46 chronic conditions over their lifetime in 282,712 individuals from the UK Biobank. By grouping individuals based on their life-history of chronic illness, we show here that risk of incident dementia can be stratified by both the type and timing of their accumulated chronic conditions. We identified several distinct clusters of multimorbidity, and their associated risks varied in an age-specific manner. Compared to low multimorbidity, cardiometabolic and neurovascular conditions acquired before 55 years were most strongly associated with dementia. Acquisition of mental health and neurovascular conditions between the ages of 55 and 70 was associated with an over two-fold increase in dementia risk compared to low multimorbidity. The age-dependent role of multimorbidity in predicting dementia risk could be used for early stratification of individuals into high and low risk groups and inform targeted prevention strategies based on a person’s prior history of chronic disease.

## Main

50 million people worldwide are diagnosed with dementia, and this number is projected to grow rapidly. The global increase in life expectancy makes dementia prevention an urgent public health priority^3^. Up to 80% of dementia patients have two or more other chronic health conditions by the time they receive their diagnosis^2^. This makes multimorbidity a potentially crucial target for optimising dementia treatment and possibly prevention on a population level.

Multimorbidity has significant impacts on patients, their families, and health systems. Individuals with multimorbidity are more likely to use health services^4,5^, have impaired functional status^6^, and are at higher risk for mortality^7^. Like dementia, multimorbidity is a global health concern that is increasingly common in older age^8^. The rising prevalence of multimorbidity has driven prominent and pressing calls to action for the medical sciences to move away from single disease focused models of treatment and research^9–11^.

However, despite the fact that dementia has consistently been associated with multimorbidity ^12,13^, there are large gaps in our understanding of this relationship. For instance, multimorbidity has most commonly been quantified as a binary classification (yes/no), or as a count of the number of pre-existing conditions^11,14^. Neither framework considers interactions between conditions, or recognizes that patterns of co-occurring conditions can cluster in a systematic, predictable fashion ^9^. Advancing beyond a simplistic measurement of multimorbidity has previously been limited by lack of large datasets with linked electronic health records. Moreover, many studies do not examine a comprehensive range of conditions; a review of 556 multimorbidity studies reported the median number of included conditions to be only 17, with only 8 conditions consistently included (diabetes, stroke, cancer, chronic obstructive pulmonary disease, hypertension, coronary heart disease, chronic kidney disease, and heart failure)^14^. Finally, the assessment of longitudinal trajectories of multimorbidity is lacking, even in those studies which include a relatively comprehensive list of conditions ^10,11^. Although multimorbidity is increasingly common in older age, it is not solely confined to late life ^8,15^. For example, in a Scottish population, nearly two-thirds of 65-84 year olds were multimorbid but almost a third of 45 - 64 year olds were also identified as multimorbid^8^. Therefore, characterising the sequential accumulation of multimorbidity patterns throughout the lifespan is a necessary step in order to bring our understanding of multimorbidity as a risk factor in line with the current life-course perspective of other dementia risk factors ^16^.

Here, we address these challenges by characterising the sequential patterns of multimorbidity throughout the life course. We use the UK Biobank, a large population cohort of over 502,000 individuals^17^. The baseline assessment was conducted in the late 2000s, when participants were 40 to 69 years old. Importantly, the UK Biobank has continuously updated linked electronic health records since the baseline assessment occurred, allowing us to study chronic conditions diagnosed prior to the baseline assessment as well as up to an average of 12 years after. To fully take advantage of this unique resource, we identify patterns of co–occurring chronic conditions (termed multimorbidity clusters) in each of three distinct age ranges: 0-55 years, 55 - 65 years, and 65-70 years. Within each age range, we apply a clustering procedure which classifies participants into distinct multimorbidity clusters based on their diagnoses within the given portion of the lifespan. This enables us to identify the patterns of chronic conditions which tend to co-occur across the study population in each age range, cluster participants in distinct fashion based on their unique accumulated conditions within each age range, and study how participants transition between different multimorbidity clusters across the lifecourse. This framework allows us to answer three key questions: 1) how do patterns of multimorbidity change during the lifespan?; 2) do different multimorbidity patterns confer different levels of risk for future dementia?; and 3) how does multimorbidity accumulate over the life-course in dementia patients? (Figure 1)

**Figure 1:**
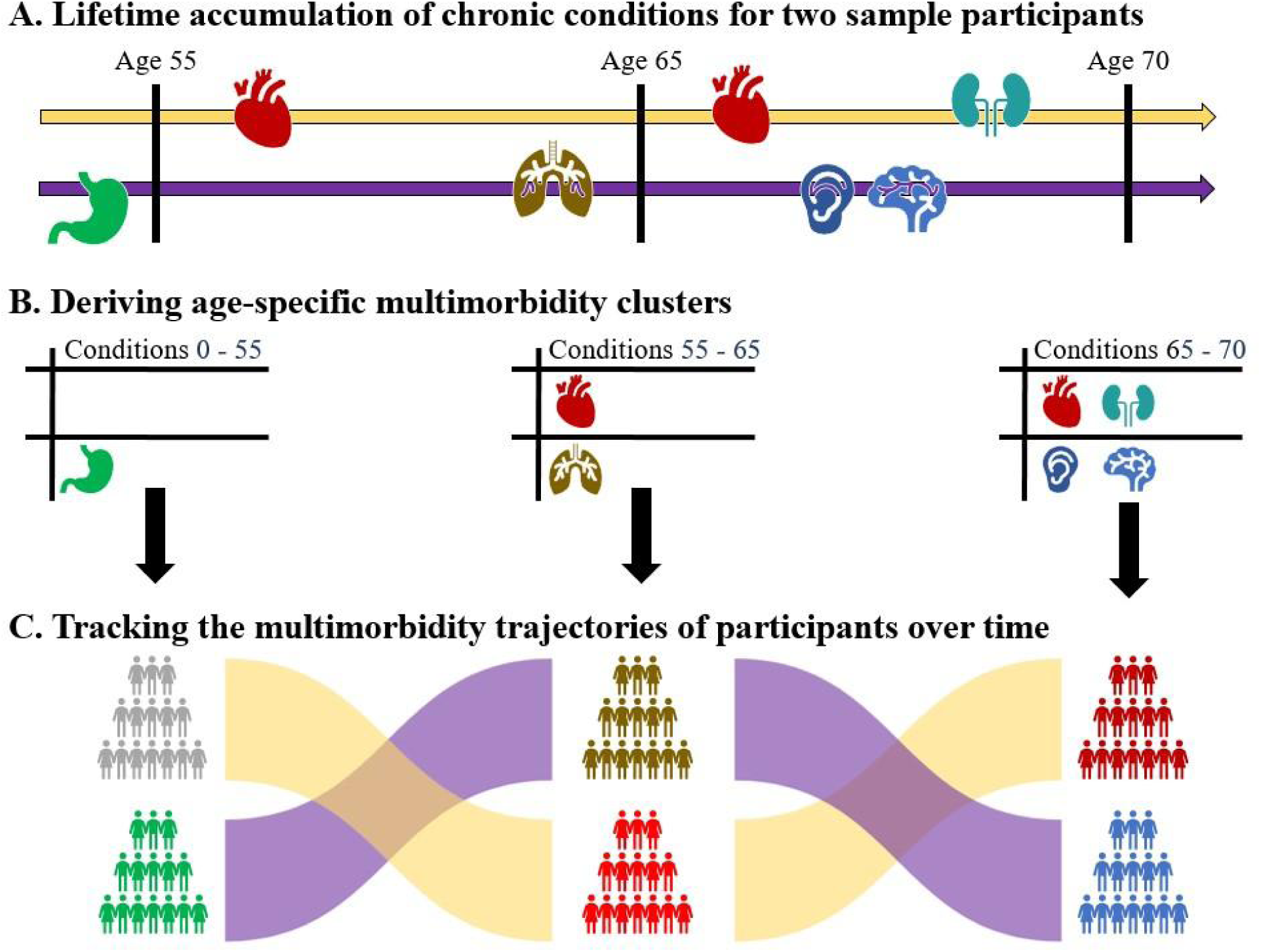
Tracking lifetime accumulation of multimorbidity: Study overview showing the derivation of lifetime multimorbidity patterns for two sample subjects. A) For each participant, we ascertain the diagnosis of a variety of chronic conditions using linked electronic health records. Based on the age of onset, diagnoses are considered in only one of three distinct age ranges: 0-55 years, 55-65 years, and 65-70 years, representing early, mid-, and late-life. B) Within each age range, a clustering analysis is performed. At age 55, participants are clustered into groups based on their conditions diagnosed from birth to age 55. At age 65, participants are clustered based on their conditions diagnosed between ages 55 and 65. At age 70, participants are clustered based on their conditions diagnosed between ages 65 and 70. C) We track the movement of participants between clusters across the different age ranges. Participants may move between clusters describing different multimorbidity patterns (eg gastrointestinal -> respiratory), or, may move between clusters defined by similar conditions (eg. cardiovascular -> cardiovascular), depending on their unique pattern of accumulated conditions.

## Results

We tracked the movement of participants across distinct life-course patterns of multimorbidity. In each of the three age ranges assessed, we found multimorbidity clusters defined by cardiometabolic, neurovascular, and mental health conditions. Additionally, in the 0-55 age range only, we also identified a cluster of eye conditions, while a peripheral vascular cluster was found in both of the later age ranges. Crucially, we not only show that dementia risk is most associated with cardiovascular, mental health, and neurological related multimorbidity, but also that the odds of developing dementia varies based on ***when*** in the lifespan these conditions are diagnosed. Prior to midlife (55 years of age), development of cardiometabolic conditions was associated with the highest risk of incident dementia. However this changed from mid-to late-life; in the 55-65 and 65-70 age ranges, the accumulation of mental health and neurovascular conditions was associated with the highest risk, a two-fold increased risk of dementia. The dementia-specific trajectories of multimorbidity identified in this study could be used for early stratification of individuals into high and low risk groups and inform targeted prevention strategies based on a person’s prior history of chronic disease.

### Participants

We used data from 282,712 UK Biobank^17^ participants with available medical records after the age of 65, who were dementia free at age 65, and who were diagnosed with at least one chronic condition other than dementia. The mean age at baseline was 61.6 (SD=4.65), and 150,687 participants were female (53.3%). After the age of 65, 6,922 (2.45%) participants had developed dementia. Table 1 displays demographic statistics stratified by incident dementia status. The majority of dementia patients (70.75%) had at least four other conditions diagnosed by the time they received a diagnosis of dementia, compared to 51.78% of controls having four or more chronic conditions diagnosed throughout the study period.

**Table 1:** Demographic characteristics of the analysis sample. Mean and standard deviation are given for age at the baseline UK biobank assessment, and years of education. For sex and number of conditions, the quantity and percentage of the sample is listed.

|  | **Controls (N = 275790)** | **Dementia (N = 6922)** |
| --- | --- | --- |
| **Age at baseline, years (SD)** | 61.51 (4.65) | 65.29 (3.24) |
| **Female (%)** | 147,409 (53.45%) | 3,278 (47.36%) |
| **Education, years (SD)** | 13.13 (3.07) | 12.40 (3.05) |
| **Number of chronic conditions (%)** |  |  |
| **1** | 46,792 (16.97%) | 496 (7.17%) |
| **2** | 45,390 (16.46%) | 749 (10.82%) |
| **3** | 40,806 (14.80%) | 780 (11.27%) |
| **>=4** | 142,802 (51.78%) | 4,897 (70.75%) |

### Age-specific patterns of multimorbidity

To estimate life-course patterns of multimorbidity, we studied 46 chronic conditions diagnosed within three distinct age ranges: 0-55 years, 55-65 years, and 65-70 years. Within each age range, we clustered participants based on new chronic conditions accumulated only within that age range (Figure 1). We employed a two-stage multivariate clustering procedure (Methods) which created clusters based on chronic conditions which co-occurred together in a non-random, systematic fashion. The resulting multimorbidity clusters delineate participants into distinct groups, each with a unique pattern of chronic conditions diagnosed within a given age range. Output clusters are named according to prevalence of, and comorbidity between, chronic conditions within the cluster relative to patterns in the full sample (Methods). Five clusters were identified as the best fit within each age range.

### Multimorbidity from 0-55 years

The five multimorbidity clusters are described as: 1) a relatively healthy group of participants with low multimorbidity burden (LOW, N=243855); 2) a group with mental health conditions (MH, N=23116); 3) a group with cardiometabolic conditions (CVMTB, N=10971); 4) a group with eye conditions (EYE, N=3302); and 5) a group with neurovascular conditions (NVASC, N=1468). The multimorbidity burden was lowest in the LOW group (mean number of chronic conditions = 0.27 +-0.57), and highest in the NVASC group (2.92 +-2.05). The MH group had a higher percentage of females (67.74%) than males, while the CVMTB (35.92%) and NVASC (41.49%) groups had a lower percentage of females (Table 2).

**Table 2:**
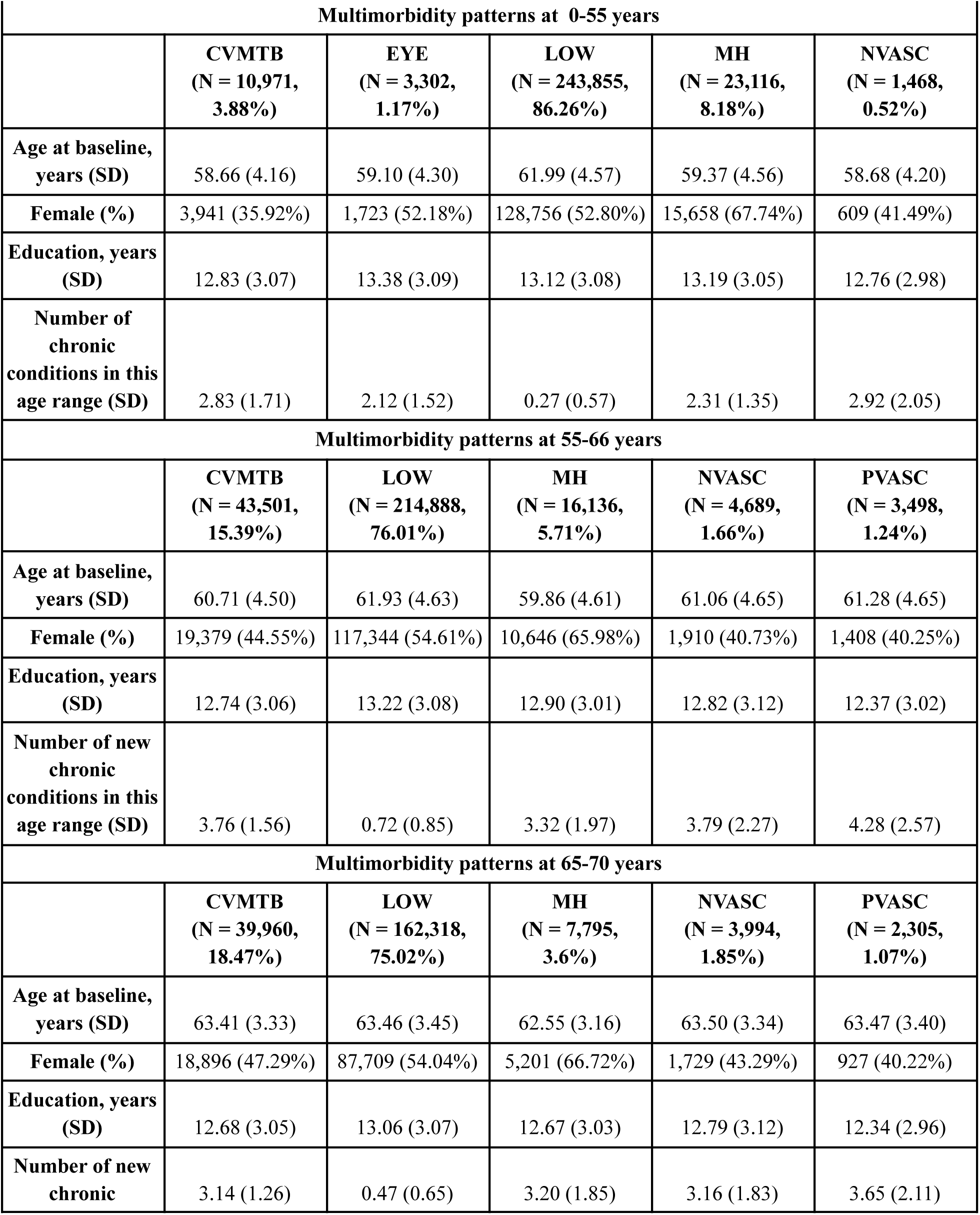
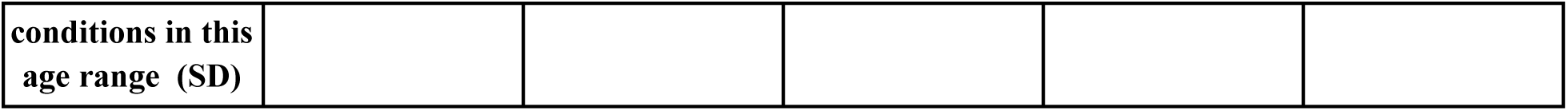
Demographic characteristics and disease burden of multimorbidity clusters. . Age at baseline, number and proportion of females, years of education, and mean (sd) number of chronic conditions are given for each multimorbidity cluster. Note that the number of chronic conditions refers only to the average number of newly acquired diagnoses received in that age range.

The observed/expected (OE) ratio (OE) is used to describe the notable conditions within each cluster. For example, an OE ratio of 2 for diabetes in a given cluster indicates that there are twice as many participants in that cluster with diabetes than expected, based on the prevalence of diabetes in the entire sample (Methods).

Comorbidity clusters are represented by chord diagrams, circular figures with nodes along the circumference and lines (edges) connecting each pair of nodes. The size of nodes is scaled to the OE of a given condition, while the edges are colour coded to show high and low comorbidity (Figures 2-4), such that orange-yellow links between a pair of conditions represents a larger than expected comorbidity (Methods).

**Figure 2:**
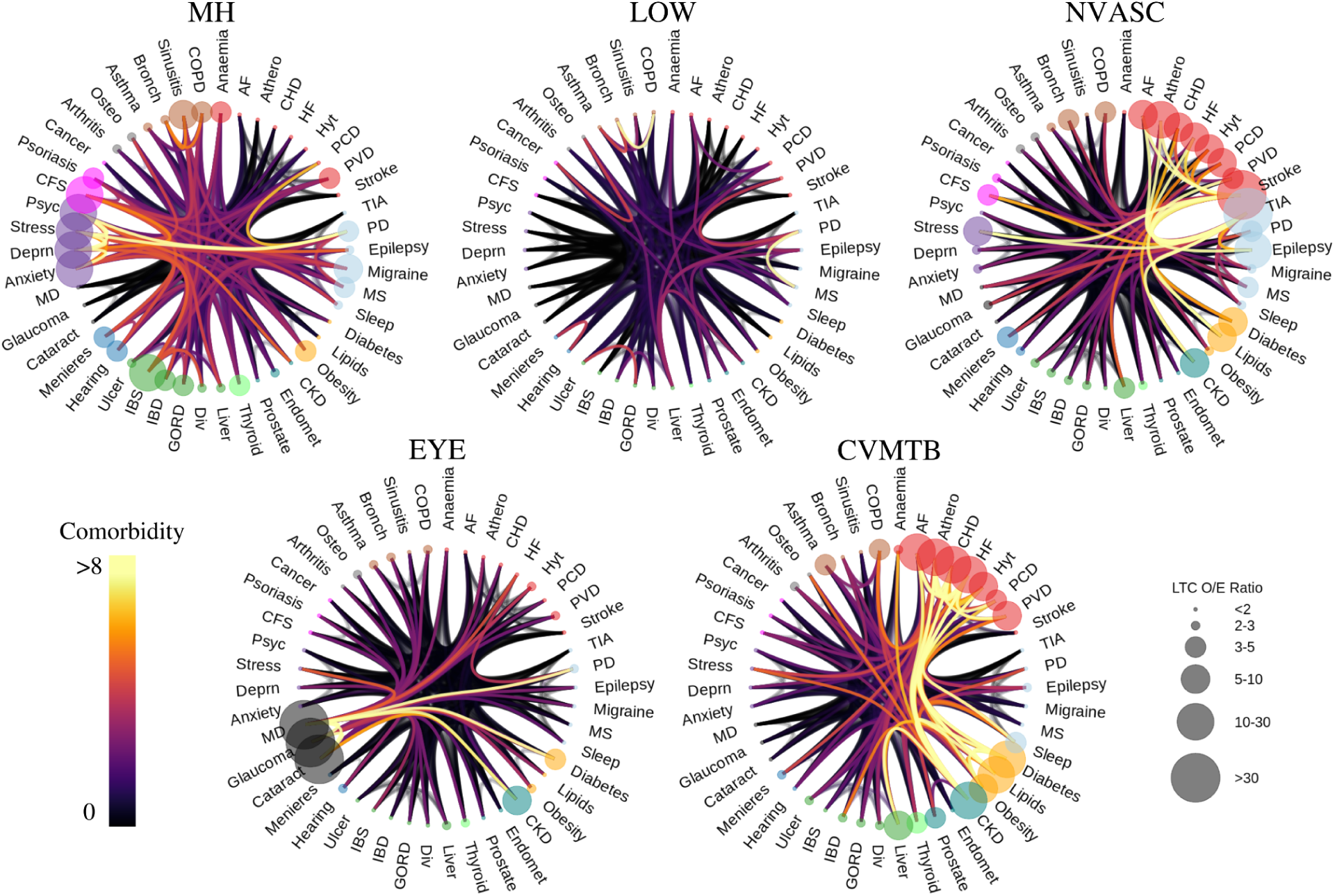
Multimorbidity patterns from birth to age 55 are defined by mental health, cardiometabolic, neurovascular, eye, and low disease burden patterns. For each multimorbidity cluster identified from age 0-55 years, we plot a chord diagram describing the defining conditions and their comorbidity patterns. Each condition is represented by a dot along the circumference of the cluster diagram. The dots are colour coded according to ICD-10 classification (eg. starting with anaemia and moving clockwise, cardiovascular conditions are in red, neurological conditions are in light blue, metabolic conditions are in yellow, genitourinary conditions are in teal, thyroid conditions are in light green, gastrointestinal conditions are in green, sensory (hearing) conditions are in blue, sensory (eye) conditions are in black, mental health conditions are in purple, chronic fatigue syndrome, psoriasis, and cancer are in pink (grouped together as ‘other’ conditions), musculoskeletal conditions are in grey, and respiratory conditions are in brown). The size of the dot is scaled to match the observed/expected (OE) ratio, such that a larger OE represents a larger than expected proportion of participants in the cluster having a diagnosis of a given chronic condition. Lines connecting pairs of conditions are colour coded to represent the strength of comorbidity between two given conditions, such that orange-yellow lines represent strong comorbidity, while black lines indicate low comorbidity. The clusters are named according to the OE and comorbidity patterns. Moving clockwise from top left, the multimorbidity clusters from birth to age 55 are a mental health cluster (MH), a low disease burden/health cluster (LOW), a neurovascular cluster (NVASC), a cardiometabolic cluster (CVMTB), and an eye cluster (EYE).

Figure 2 illustrates each cluster within the 0-55 age range. As expected, no condition is overrepresented in the LOW group, suggesting low/no multimorbidity in this group. The CVMTB group is so named because of an over-representation of cardiovascular and metabolic conditions in this cluster. This group also includes a higher than expected number of individuals with chronic kidney disease, liver disease, and respiratory conditions, although these were less prevalent than the cardio-metabolic conditions. The EYE group is dominated by the co-occurence of eye conditions such as macular degeneration, glaucoma, and cataract, but also contains a higher than expected number of individuals with chronic kidney disease and diabetes.

The NVASC group is dominated by stroke and TIA, but also includes a larger than expected proportion of individuals with other neurological conditions, cardiovascular conditions, stress disorders, and metabolic disorders. The MH group is defined by the over-representation of mental health conditions (psychoses, anxiety disorders, depression, and stress disorders). There was also a larger than expected proportion of chronic fatigue syndrome, psoriasis, gastrointestinal conditions (irritable bowel syndrome, inflammatory bowel disease), neurological conditions (epilepsy, multiple sclerosis, Parkinson’s disease), and respiratory conditions in the MH group.

### Multimorbidity from 55-65 years

The five multimorbidity clusters are described as: 1) a relatively healthy group of participants (LOW, N=214888); 2) a group with mental health conditions (MH, N=16136); 3) a group with cardiometabolic conditions (CVMTB, N=43501); 4) a group with peripheral vascular conditions (PVASC, N=3498); and 5) a group with neurovascular conditions (NVASC, N=4689). The multimorbidity burden was lowest in the LOW group (mean number of chronic conditions = 0.72 +-0.85), and highest in the PVASC group (4.28 +-2.57).

The MH group had a higher percentage of females (65.98%), while the CVMTB (44.55%), NVASC (40.73%), and PVASC (40.25%) groups had a lower percentage of females than males. (Table 2). Compared to the multimorbidity patterns in the 0-55 age range, the 55-65 range had fewer participants in the LOW and MH groups, and more participants in the CVMTB group. While disease burden in the LOW group remained similar as in the younger age range, in all other groups participants developed, on average, over three new chronic conditions between the ages of 55 and 65 (Table 2).

Figure 3 plots the OE patterns of each cluster. As expected, no condition is over-represented in the LOW group. The CVMTB group has an over-representation of cardiovascular and metabolic conditions and also a higher than expected number of individuals with chronic kidney disease, liver disease, and chronic obstructive pulmonary disorder. The NVASC group is again dominated by stroke and TIA, but also includes a larger than expected proportion of individuals with other neurological conditions (epilepsy, migraine), cardiovascular conditions (heart failure, atrial fibrillation), chronic fatigue syndrome, lipid disorders, chronic kidney disease, and stress disorders. The MH group has an over-representation of mental health conditions (psychoses, anxiety disorders, depression, and stress disorders), but there was also a larger than expected proportion of chronic fatigue syndrome, psoriasis, and neurological conditions (epilepsy, migraine, Parkinson’s disease). In the PVASC group, which did not appear in the 0-55 age range, the most prominent conditions are atherosclerosis and other peripheral vascular disorders. Other conditions over represented include cardiovascular conditions (heart failure, CHD, atrial fibrillation), metabolic conditions, chronic kidney diseases, depression, and chronic obstructive pulmonary disorder. The EYE group from 0-55 years did not appear in the 55-65 year age range.

**Figure 3:**
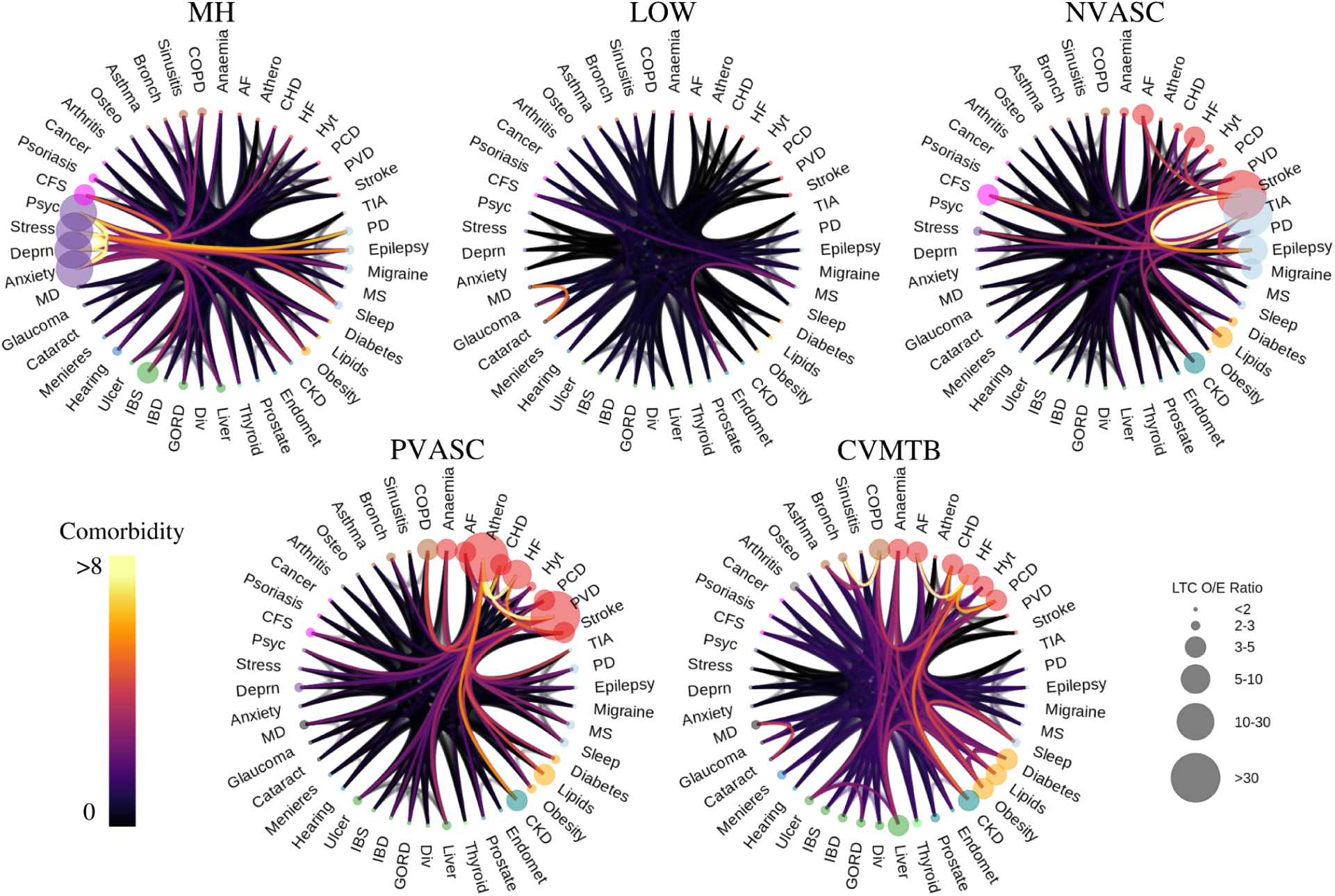
Multimorbidity patterns from age 55-65 are defined by mental health, cardiometabolic, neurovascular, peripheral vascular, and low disease burden patterns. Figure 2 plots, for each multimorbidity cluster identified from age 55-65 years, a chord diagram describing the defining conditions and their comorbidity patterns. Moving clockwise from top left, the multimorbidity clusters from age 55-65 are a mental health cluster (MH), a low disease burden/health cluster (LOW), a neurovascular cluster (NVASC), a cardiometabolic cluster (CVMTB), and a peripheral vascular (PVASC).

### Multimorbidity from 65 - 70 years

We identified five multimorbidity clusters, similar to the 55-65 age range : 1) LOW N=162318; 2) MH, N=7795; 3) CVMTB, N=39960), defined by an over-representation of cardiovascular (except atherosclerosis) and metabolic conditions and a higher than expected number of individuals with chronic kidney disease, endometriosis, gastrointestinal conditions, respiratory conditions, and sleep disorders. ; 4) PVASC (N=2305),; and 5) NVASC (N=3994), dominated by stroke and TIA, but also including a larger than expected proportion of individuals with other neurological conditions (epilepsy, migraine), cardiovascular conditions, chronic fatigue syndrome, lipid disorders, chronic kidney disease, and chronic obstructive pulmonary disorder.. As in the 55-65 range, the multimorbidity burden was lowest in the LOW group had the lowest multimorbidity burden (mean number of chronic conditions = 0.47 +-0.65), and highest in the PVASC group (3.65 +-2.11)

The MH group had more females (66.72%), while the CVMTB (47.29%), NVASC (43.29%), and PVASC (40.22%) groups had fewer females than males. (Table 2). There were fewer participants in the LOW and MH groups at 65-70 than at 55-65, despite the shorter age range. Numbers in the CVMTB, PVASC, and NVASC groups remained relatively stable across the ages. While disease burden in the LOW group remained similar, in all other groups participants developed, on average, over three new chronic conditions between the ages of 65 and 70 (Table 2). 7,465 individuals who had a death record between ages 65-70, 1102 who received a diagnosis of dementia between ages 65-70, and 57,773 individuals who did not have health records past age 70 were not included in the multimorbidity clustering procedure in the 65-70 age range. 216,372 were included in this analysis (Methods).

Figure 4 plots the OE patterns of each cluster, which largely shows similar characteristics as clusters in the 55-65 age range. Thus, the same naming conventions are used for clusters of conditions developed between age 65-70.

**Figure 4:**
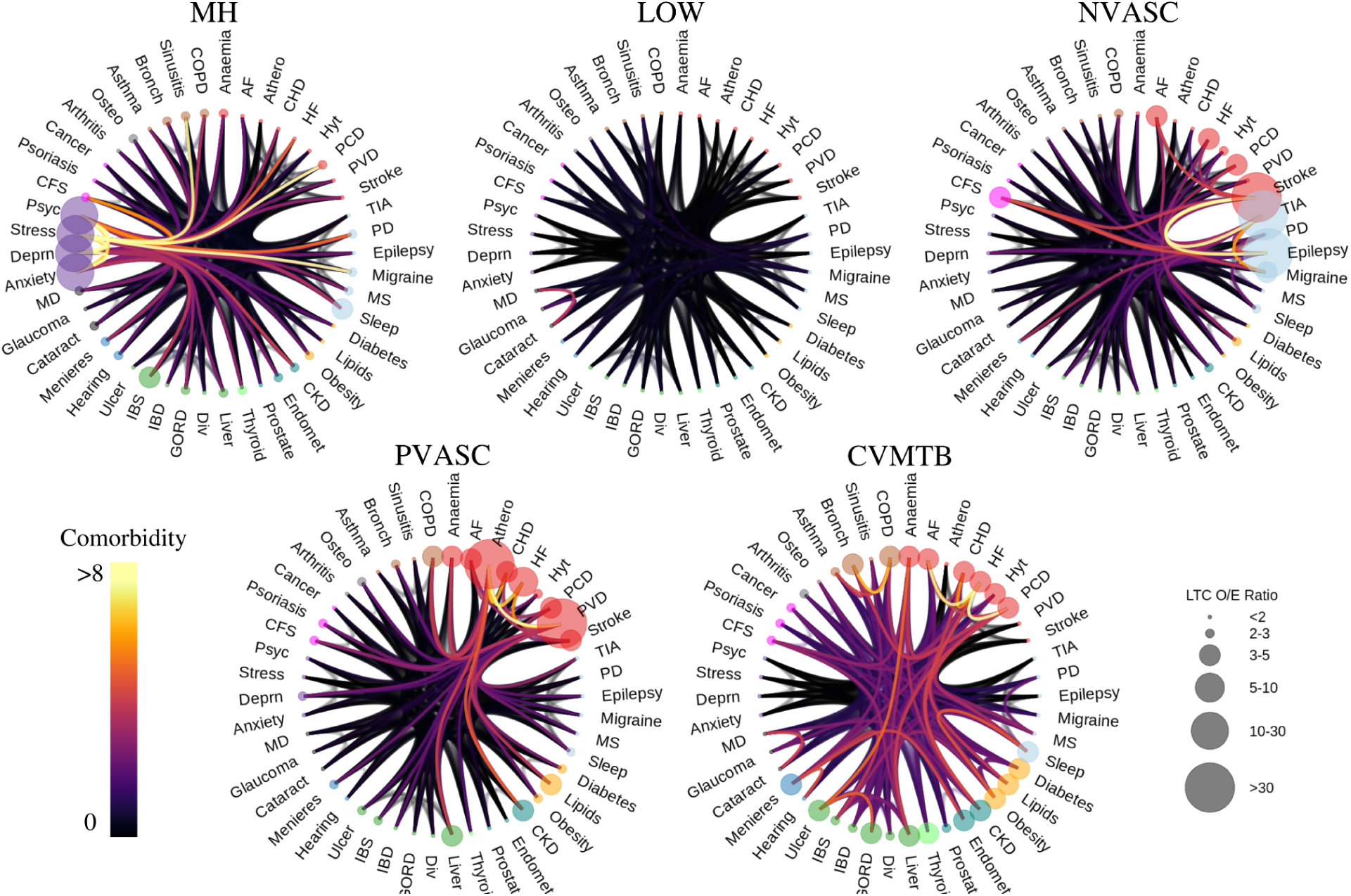
Multimorbidity patterns from age 65-70 are defined by mental health, cardiometabolic, neurovascular, peripheral vascular, and low disease burden patterns. Figure 2 plots, for each multimorbidity cluster identified from age 65-70 years, a chord diagram describing the defining conditions and their comorbidity patterns. Moving clockwise from top left, the multimorbidity clusters from age 65-70 are a mental health cluster (MH), a low disease burden/health cluster (LOW), a neurovascular cluster (NVASC), a cardiometabolic cluster (CVMTB), and a peripheral vascular (PVASC).

### Multimorbidity Clusters associated with dementia

To assess the association of multimorbidity clusters with incident dementia, we used a logistic regression with incident dementia as outcome, multimorbidity cluster as predictor, and age, sex, and years of education as covariates. The LOW group was used as the reference group. To first assess the cross-sectional association of each multimorbidity cluster, irrespective of prior or future multimorbidity, we analysed each age range independently and found all multimorbidity clusters were significantly associated with increased risk of incident dementia (𝑃_𝐹𝐷𝑅_ < 0.05) with the exception of the EYE cluster in the 0-55 age range (Methods). To assess how the accumulation of multimorbidity over the lifespan is associated with incident dementia, we instead performed a logistic regression with incident dementia as outcome, multimorbidity cluster from 0-55, multimorbidity cluster from 55-65, and multimorbidity cluster from 65-70 as predictors, and age, sex, and years of education as covariates. This identifies the risk associated with multimorbidity patterns at each time point while covarying for a participant’s past and future multimorbidity. This analysis identified both disease- and time-specific patterns of multimorbidity associated with increased risk.

In the 0-55 age range, the NVASC (OR = 1.58 95% CI = [1.11, 2.19]) and CVMTB (OR = 1.51, CI = [1.31, 1.72]) clusters were associated with increased dementia risk in comparison to the LOW cluster. At this age, belonging to the MH or EYE cluster was not associated with increased risk of dementia when compared to having low/no multimorbidity. However, this pattern was different in the 55-65 age range, where relative to the LOW cluster, the NVASC (OR = 1.84, [1.58, 2.14]), MH (OR = 1.72 [1.55, 1.9]), CVMTB (OR = 1.5 [1.4, 1.6]), and PVASC (OR=1.4, [1.16, 1.69]) clusters were associated with increased dementia risk. In the 65-70 age range, the MH (OR = 2.48 [2.21, 2.78]) and NVASC (OR = 2.46, [2.13, 2.83]) clusters stood out as having over a two-fold increase in the odds of developing dementia in comparison to the LOW cluster. The CVMTB (OR = 1.22 [1.14, 1.31]) cluster also showed increased risk in this time frame. Table 3 displays the OR of each multimorbidity cluster across the age ranges investigated.

**Table 3:** Multimorbidity impact on risk of incident dementia varies across the life course. Table 3 displays odds ratios (OR) of each multimorbidity group at each age range, representing the risk associated with each group while co-varying for a participant’s age, sex, years of education, and life course patterns of multimorbidity. 95% confidence intervals and statistical significance of the computed ORs is also shown.

| Multimorbidity patterns at 0-55 years |  |  |  |  |
| --- | --- | --- | --- | --- |
| Predictor | beta | Odds Ratio [95% CI] | *p* | *p<sub>BH</sub>* |
| Intercept | -17.09 |  | 0 | 0 |
| CVMTB | 0.61 | 1.84 [1.61, 2.1] | 1.43E-19 | 2.86E-19 |
| EYE | 0.06 | 1.06 [0.78, 1.4] | 0.69 | 0.69 |
| MH | 0.12 | 1.13 [1.02, 1.26] | 0.02 | 0.03 |
| NVASC | 0.67 | 1.96 [1.37, 2.71] | 9.33E-05 | 1.24E-04 |
| Age | 0.22 | 1.24 [1.24, 1.25] | 0 | 0 |
| Male Sex | 0.16 | 1.18 [1.12, 1.24] | 7.49E-11 | 1.20E-10 |
| Education (years) | -0.05 | 0.95 [0.95, 0.96] | 1.88E-27 | 5.02E-27 |
| Multimorbidity patterns at 55-65 years |  |  |  |  |
| Predictor | beta | Odds Ratio [95% CI] | *p* | *p<sub>BH</sub>* |
| Intercept | -17.51 |  | 0 | 0 |
| CVMTB | 0.49 | 1.64 [1.53, 1.75] | 2.06E-49 | 5.50E-49 |
| EYE | 0.63 | 1.88 [1.7, 2.08] | 2.83E-34 | 5.66E-34 |
| NVASC | 0.77 | 2.16 [1.85, 2.5] | 3.10E-23 | 4.95E-23 |
| PVASC | 0.55 | 1.73 [1.43, 2.08] | 8.97E-09 | 8.97E-09 |
| Age | 0.22 | 1.25 [1.24, 1.26] | 0 | 0 |
| Male Sex | 0.15 | 1.16 [1.1, 1.22] | 4.33E-09 | 4.95E-09 |
| Education (years) | -0.04 | 0.96 [0.95, 0.97] | 5.75E-22 | 7.67E-22 |
| Multimorbidity patterns at 65-70 years |  |  |  |  |
| Predictor | beta | Odds Ratio [95% CI] | *p* | *p<sub>BH</sub>* |
| Intercept | -19.84 |  | 0 | 0 |
| CVMTB | 0.21 | 1.24 [1.16, 1.33] | 5.21E-10 | 6.94E-10 |
| EYE | 0.93 | 2.54 [2.26, 2.84] | 2.66E-56 | 7.10E-56 |
| NVASC | 0.96 | 2.61 [2.27, 2.99] | 8.87E-42 | 1.77E-41 |
| PVASC | 0.33 | 1.39 [1.09, 1.73] | 0.01 | 0.01 |
| Age | 0.26 | 1.29 [1.28, 1.3] | 0 | 0 |
| Male Sex | 0.14 | 1.15 [1.09, 1.22] | 2.75E-07 | 3.14E-07 |
| Education (years) | -0.04 | 0.96 [0.95, 0.97] | 1.19E-20 | 1.91E-20 |

### Multimorbidity Trajectories associated with dementia

Having identified the differential association of multimorbidity phenotypes with dementia at each distinct age-range, we sought to characterise the trajectory of accumulation of multimorbidity over the life-course in dementia patients. Here we use a sankey diagram to visualise the movement of dementia patients between multimorbidity categories prior to receiving a dementia diagnosis (Figure 5).

**Figure 5:**
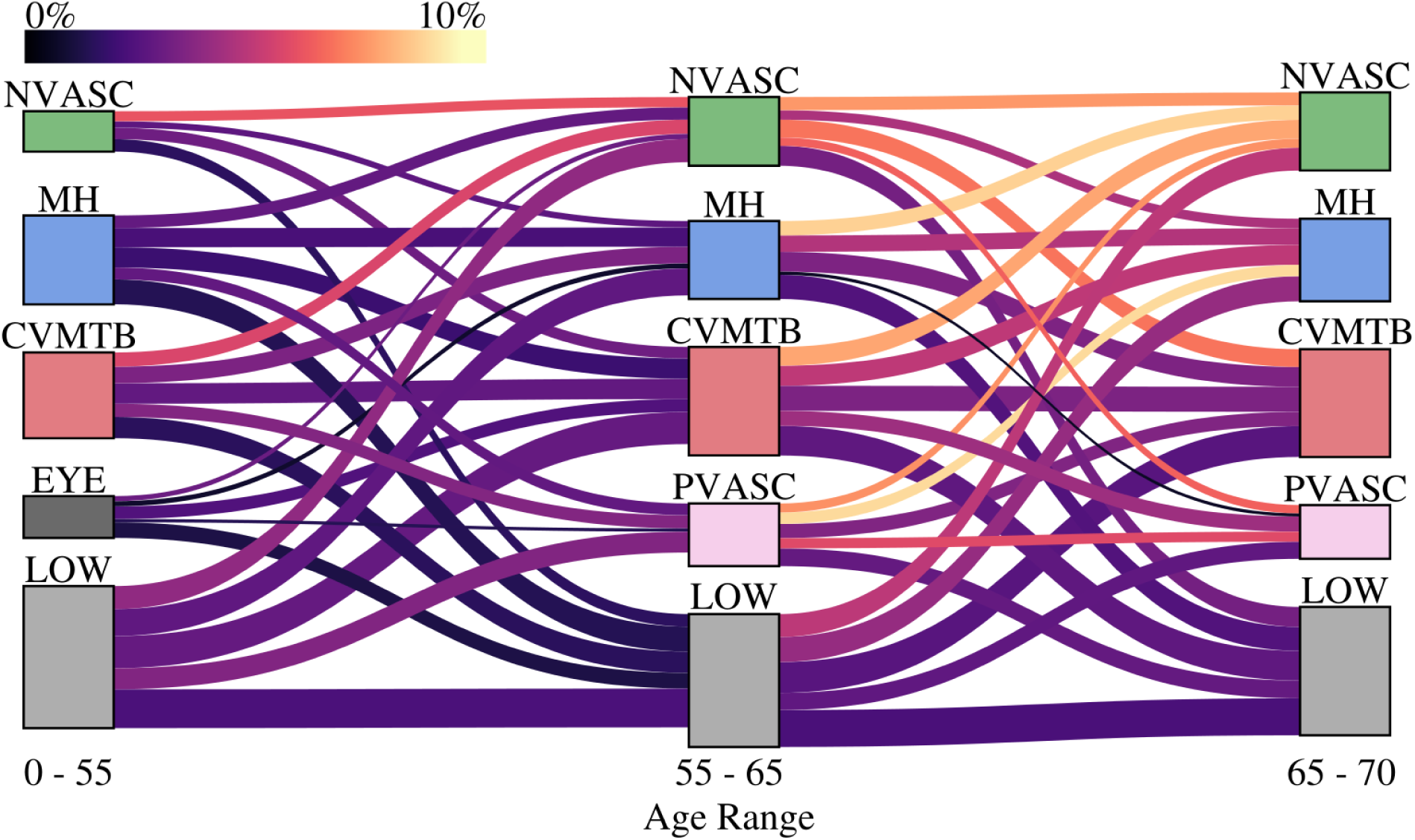
Multimorbidity trajectories of dementia patients. Our clustering analysis separates participants into groups in each of three age ranges assessed. Here, a sankey diagram is used to visualise the movement of dementia patients between multimorbidity clusters from 55 to 65, and from 65 to 70. Parameters of the figure (block height, link width, link colour) are coordinated to highlight the multimorbidity trajectories with larger proportions of participants who received a future diagnosis of dementia. The height of each block is scaled to the log of the total number of future dementia cases in a given cluster (taller blocks represent larger numbers of future patients). Similarly, the height of each link is scaled to the log of the total number of future dementia cases who follow a given trajectory. To highlight the trajectories which occur more frequently in future dementia patients, the colour of the links is scaled to the proportion of participants in the full sample following a trajectory who receive a future diagnosis of dementia. For example, the wide link between the LOW clusters at 55 and 65 indicate a relatively larger number of participants in the full sample following this trajectory who receive a future diagnosis. However, the dark purple colour of this link conveys that a relatively lower percentage (2.38%) of individuals in the full sample following this trajectory receive a future diagnosis of dementia. Conversely, the bright link between CVMTB at 65 and NVASC at 70 indicates a higher percentage (8.13%) of participants in the full sample following this trajectory receive a future diagnosis of dementia, though the overall number of individuals following this path is not as large as the LOW at 55 to LOW at 65 path.

Visualisation of the multimorbidity trajectories of dementia patients allows us to assess differential risk based on a participant’s unique pattern of accumulation of chronic conditions over their lifespan. Several notable observations can be made. Trajectories on the left side of Figure 5 generally represent lower dementia prevalence than those on the right side, indicating that delaying or reducing multimorbidity burden until the age of 65 could have key implications for risk reduction. There was a higher prevalence of incident dementia in all pathways leading to NVASC membership in the 65-70 age range, highlighting the significant risk associated with diagnosis of conditions such as stroke, TIA, and epilepsy between the ages of 65 and 70. In specific cases, the risk associated with membership in a multimorbidity cluster at a given time varied based on which diseases a person accumulated over the ***next*** five or ten years. For example, while the CVMTB group from 0-55 years is associated with increased dementia risk, this risk is most prominent for the individuals in the CVMTB group who progress to the NVASC group over the next ten years of their lives. Conversely, those in the CVMTB group at 55 who had minimal disease burden (LOW) in the next ten years had a lower risk of dementia. Interestingly, the CVMTB-NVASC pathway contained a higher proportion of incident dementia cases than the CVMTB-CVMTB pathway. Progression to the NVASC cluster at 70, as opposed to progression to any other cluster, was also associated with higher risk for participants who were in the NVASC cluster at 55 and 65, the MH cluster at 65, and the CVMTB cluster at 65. Other trajectories notably containing high prevalence of incident dementia include PVASC at 65 to MH at 70 and NVASC at 65 to either CVMTB or PVASC at 70. Throughout the lifespan, progression to membership in the LOW cluster was associated with a significantly lower risk in comparison to any other trajectory.

## Discussion

Identification of at-risk individuals is essential for any successful modification of the disease course of dementia. We clustered participants based on accumulation of chronic conditions within three distinct age ranges, and present three novel and salient suggestions which help bring understanding of multimorbidity as a risk factor in line with the current life-course perspective of other dementia risk factors^16^. First, we note that the link between multimorbidity and increased risk of incident dementia can be stratified by both the ***type*** and ***timing*** of their accumulated chronic conditions.

Second, we find that participants who repeatedly accumulated chronic conditions throughout their lifespan had significantly higher odds of dementia, whereas in those who remained in the “low” cluster until age 65, and who moved from a multimorbidity cluster to the “low” cluster at the next time point, odds of developing dementia was considerably lower. This suggests that delaying the development of a new chronic condition before the age of 65 could have significant implications for dementia risk reduction for all individuals, regardless of the diseases they accumulated prior to age 55.

Third, we found that a diagnosis of either cardiometabolic or neurovascular conditions by the age of 55 was associated with increased risk of future dementia relative to low/no multimorbidity. Developing a new cardiometabolic, neurovascular, mental health, or peripheral vascular condition between the ages of 55 and 65 was also associated with higher odds of dementia. In this critical age range, compared to the multimorbidity-dementia link at age 55, the risk associated with cardiometabolic conditions remained relatively stable while the risk associated with new mental health and neurovascular conditions increased. This trend continued in the 65-70 age range, in which the accumulation of either mental health or neurovascular conditions was associated with an over two-fold increase in the odds of future dementia. Development of cardiometabolic conditions remained associated with increased risk, although to a lesser degree than previous age ranges.

Further, we present the unique ***trajectories*** of multimorbidity that were most strongly associated with dementia. From age 55 until 65, the NVASC-NVASC, CVMTB-NVASC, and LOW-NVASC trajectories were most strongly associated with future dementia, with 6.3%, 5.9%, and 4.1% of all participants following these trajectories going on to develop dementia, respectively. This highlights the importance of delaying onset of neurovascular and cardiometabolic conditions until age 65 for reducing dementia risk. Between age 65 and 70, the PVASC-MH, MH-NVASC, and CVMTB-NVASC trajectories were most strongly associated with future dementia, with 9.2%, 9.1%, and 8.1% of all participants following these trajectories going on to develop dementia, respectively. This highlights the emergence of mental health conditions between ages 65 and 70 as strong risk factors for dementia, as well as the continued risk associated with neurovascular conditions.

To date, few studies have assessed either trajectories^18^ or clusters^19,20^ of multimorbidity in the context of dementia risk, and no study has assessed how the longitudinal accumulation of chronic conditions influences dementia risk. A large study using Danish registry data computed temporal trajectories of individual chronic conditions prior to a diagnosis of Alzheimer’s disease and compared them to trajectories preceding vascular dementia^18^. Diabetes, hypertension, heart failure, depression, intracerebral haemorrhage, and atherosclerosis were among the conditions identified as commonly occurring prior to an AD or VaD diagnosis. While this study looked at accumulation of multimorbidity, it did not consider age of onset or multimorbidity clusters. Our findings add vital new information on how risk varies based on ***when*** and ***in what combination*** the above conditions are diagnosed.

Two recent studies have studied multimorbidity clusters in the context of dementia risk. In one, a mental health cluster and a cardiometabolic cluster were most strongly associated with dementia risk. An inflammatory/autoimmune cluster centred on arthritis and psoriasis was also moderately linked to higher risk^20^. In the second study, a sex stratified analysis was performed. In women, a hypertension, diabetes, and CHD cluster, along with an osteoporosis and dyspepsia cluster were associated with highest risk. In men, a diabetes and hypertension cluster, along with a CHD, hypertension, and stroke cluster were associated with highest risk ^19^. Both these studies analysed the UK Biobank population, though neither considered the longitudinal progression of multimorbidity. These findings and ours add to overwhelmingly strong evidence linking cardiometabolic (eg. CHD, hypertension, diabetes, lipid disorders), neurovascular (e.g. stroke), and mental health (e.g. schizophrenia, depression, anxiety) conditions to increased risk of dementia.

These studies both describe clusters with an inflammatory/immune component (e.g. conditions such as arthritis, psoriasis, and osteoporosis), as being linked to higher risk of dementia. While none of the clusters in our study were dominated by the prevalence or comorbidity of these conditions, arthritis, psoriasis, and osteoporosis did occur in greater than expected proportions (i.e. OE > 2) in certain clusters, particularly in the 0-55 age range. Thus, we also find some evidence of inflammatory/immune conditions contributing to dementia. However, given that neither of the above studies considered the sequential progression of multimorbidity, it is possible that the higher risk attributable to these conditions would be masked, or reduced, by the development of cardiometabolic, neurovascular, or mental health conditions in the future and not considered in their cross-sectional analyses.

### Strengths

The primary strength of our study is the integration of a clustering and longitudinal approach to advance understanding of multimorbidity as a risk factor in line with the current life-course perspective of other dementia risk factors. To derive multimorbidity clusters we used MCA, a dimensionality reduction approach which strives to identify pairs of conditions which co-occur in greater proportions than is expected based on their prevalence in the sample. This gives our study the ability to identify strong comorbidity patterns between conditions even if they have a lower overall prevalence. This is crucial for identifying mental health clusters which have proven to be a common finding in studies of multimorbidity patterns ^2,6,21,22^, but may be hard to detect if the prevalence of mental health conditions is lower in the sample ^19^. We also employed a longitudinal approach by delineating electronic health record derived diagnoses into specific age ranges, allowing us to identify critical periods in which multimorbidity conveys higher or lower risk, along with the differential risk associated with certain clusters. The use of age ranges, as opposed to utilising exact ages, both accounts for uncertainty associated with diagnosis dates in electronic health records, and eliminates methodological issues arising when multiple diagnoses are linked to the same date.

### Limitations

Four key limitations require consideration in this study. First, the UK Biobank lacks a gold standard, clinically adjudicated diagnosis of dementia. However our approach, combining hospital records, death records, and primary care data, has demonstrated a positive predictive value of 82.5% compared to clinical adjudication^23^. Combined with the size of the dataset, this makes UK Biobank a unique resource to study dementia outcomes. The positive predictive value of dementia subtypes was reported to be lower, and thus we only consider all-cause dementia as opposed to Alzheimer’s Disease, vascular dementia, or other subtypes. Second, the UK Biobank cohort is predominantly Caucasian, and has a lower incidence of chronic conditions than the general population^24^. As dementia risk and onset varies by race and ethnicity^25^, further evaluation in more diverse cohorts is required before large generalisation of our findings. Third, the observational nature of this cohort and long prodrome of dementia precludes us from making any inferences regarding causation. We also note that the presence of multimorbidity may increase the likelihood of an individual seeking medical attention and further compounding the number of diagnoses received. Fourth, we did not consider treatments being administered to participants (eg. medication). While we focus on age of diagnosis, we did not have access to other continually updated measures of disease severity. Thus, our findings remain associative in nature.

## Conclusion

We studied the accumulation of multimorbidity throughout the life course in a large prospective cohort and found that the strong association between multimorbidity and a future diagnosis of dementia varies based on the specific type of conditions an individual has, as well as when they received a diagnosis. Until midlife (age 55), the accumulation of cardiometabolic conditions, such as coronary heart disease, atrial fibrillation, heart failure, diabetes, and lipid disorders, was strongly associated with dementia risk. However from ages 55 to 70, the accumulation of mental health conditions, such as anxiety, depression, psychoses, and stress, as well as neurovascular conditions, such as stroke, transient ischaemic attack, and epilepsy, was most strongly linked to dementia. Crucially, individuals who continuously and sequentially accumulate cardiometabolic, mental health, and neurovascular conditions were at greatest risk. These findings promote the importance of considering multimorbidity patterns across the life course when assessing an individual’s risk for dementia.

## Methods

### Participants

We used data from the UK Biobank, a large population study of 502,386 individuals ^17^. Assessments were conducted at one of 22 centres throughout Scotland, England, and Wales. At the baseline assessment conducted between 2006-2010, participants were aged between 40-69 years old. Health outcomes are available via linked medical records, enabling the ascertainment of dementia and other chronic conditions throughout the lifespan, prior to and after the baseline assessment. To reduce the likelihood of including monogenetic cases of early-onset dementia^13^, we excluded any individuals for whom we could not confirm dementia free status at the age of 65. Thus, we excluded any individuals who did not have available medical records after the age 65 (N=158,793) and any individuals who received a diagnosis of dementia (N=859) or had a death record prior to age 65 (N=4460). We also excluded any individuals who had zero diagnoses of chronic conditions (55,562) in order to focus our analysis on patterns of multimorbidity. This produced an analysis sample of 282,712 individuals. The UK Biobank study received ethical approval from the Northwest Multi-Centre Research Ethics Committee. All participants provided their informed written consent.

### Ascertainment of chronic conditions

Ascertainment of chronic condition diagnoses was completed using three complementary sources: hospital inpatient data, primary care data, and death records. Hospital inpatient data was available for the full cohort, and describes the date of admission and diagnoses using International Classification of diseases, Tenth Version (ICD-10) coding. Primary care data was available for 230,000 participants, and includes coded diagnoses and dates from GP systems via READ2 and CTV-3 codes. Data on the date and cause of death was available via linkage to national death registries, coded using ICD-10 codes. Hospital and death records were available up to October 1st, 2021. Primary care data was available up to 2017.

We developed a list of 46 chronic conditions to include based on previous recommendations^21^ and a survey of the multimorbidity literature. For each, we parsed the above sources to ascertain if and when an individual received a diagnosis. If an individual received multiple diagnoses of the same condition (via repetition in the same data source or across multiple sources), the date of diagnosis was taken as the earliest available record. Importantly, diagnoses of chronic conditions were only considered if they occurred *prior to* a dementia diagnosis. The age at which a condition was diagnosed was then computed by comparing diagnosis date to the participant’s date of birth. We had access to year and month of birth, and so the midpoint of the birth month was taken as the date of birth for all participants. The ICD-10, READ2, and CTV-3 codes used to identify each condition were developed via merging of related works^26,27^ as well UK Biobank guidelines (https://biobank.ndph.ox.ac.uk/showcase/refer.cgi?id=592). Codings are listed at https://git.fmrib.ox.ac.uk/psp365/mm_risk/-/blob/main/ltc_codes.csv?ref_type=heads.

### Dementia Ascertainment

All-cause dementia status was ascertained based on a combination of hospital inpatient data, primary care data, and death report data as done in several papers based on this cohort^28,29^. To increase diagnostic accuracy, we excluded 117 individuals who self-reported having dementia but had no corresponding medical record. The list of ICD-10 codes are available at https://git.fmrib.ox.ac.uk/psp365/mm_risk/-/blob/main/ltc_codes.csv?ref_type=heads.

### Multimorbidity Clusters

Multimorbidity clusters were derived using a two step procedure. First, a matrix with dimensions 282712 x 46 (number of participants x number of chronic conditions) was derived in which each matrix element was a binary (yes/no) categorization of disease diagnosis. This matrix was submitted to multiple correspondence analysis (MCA), a multivariate technique used for identifying patterns in categorical data ^30,31^ and previously used to study multimorbidity patterns^32^. Commonly described as a version of principal component analysis for categorical data, MCA maps input data to a multidimensional space by applying correspondence analysis to an indicator matrix. MCA provides components and factor scores for both participants and conditions which describe their loading within each component. Crucially, associations between categorical variables is weighted by the ^2^ statistic ^31^. Thus, output MCA components are driven by pairs of chronic conditions for which the observed proportion of individuals with both conditions is significantly different from the expected number based on overall sample prevalence. After MCA analysis, participant factor scores were inputted to k-means clustering to identify clusters of individuals loading similarly onto MCA components. The resulting groups of individuals thus share similar patterns of comorbid chronic conditions (i.e. multimorbidity patterns). The number of clusters was selected via the silhouette and bayesian information criterion as goodness of fit statistics.

In defining multimorbidity clusters, we note key considerations. First, the above workflow was separately applied in three distinct age ranges: 0-55 years, 55-65 years, and 65-70 years old. For each age-specific analysis, we consider diagnoses only occurring within the given time frame. For example, if an individual was diagnosed with diabetes at age 60, the matrix element corresponding to their diabetes diagnosis would be 0 (no) in both the 0-55 and 65-70 years, but 1 (yes) in the 55-65 analysis. **Second, all participants are included in each age-range analysis**. That is, an individual with linked health record data up to the age of 72 would be included in each of the three age-clustering workflows, though their input data would vary at each age, depending on when they received a diagnosis of a particular condition. Third, while clustering in the 65-70 age range, we excluded individuals who had a death record between ages 65-70, received a diagnosis of dementia between ages 65-70, or who did not have linked health records past the age of 70. Fourth, for each analysis described below, a group representing low multimorbidity load (i.e. a relatively healthier group) was used as the reference group for statistical comparisons.

### Describing Clusters

Each cluster was characterised and named according to the prevalence and patterns of chronic conditions within the cluster. Specifically, for each condition and within each cluster we computed the observed/expected ratio (OE) as the prevalence of a condition within a cluster divided by the prevalence of a condition in the entire sample. For example, an OE of 2 for diabetes in a particular cluster indicates that the prevalence of diabetes in that cluster is double the prevalence of diabetes in the sample within the age range analysed. As MCA is weighted towards identifying strong comorbidity, we quantified and considered comorbidity of each pair of conditions on a per cluster basis. Specifically, a contingency table is formed for a pair of conditions, from which the ^2^ statistic is computed. This is repeated for each pair of conditions within each cluster. The magnitude of the resulting^2^ statistic indicates the strength of comorbidity between a pair of conditions, based on the observed number of individuals diagnosed with each condition. Comorbidity clusters are represented by chord diagrams, circular figures with nodes along the circumference and lines (edges) connecting each pair of nodes. The size of nodes is scaled to the OE of a given condition, while the edges are colour coded to show high and low comorbidity (Figures 2-4). ^2^ statistics are capped at a maximum of 8 based on the skewed (right tail) distribution of comorbidity metrics across all condition-condition pairs. Thus, the colour scheme does not distinguish between pairs of conditions with comorbidity >8 and any such pairing will be viewed as having equally strong comorbidity.

### Statistical Analysis

We performed a logistic regression to test the association between multimorbidity clusters across the lifespan and risk of incident dementia. The clustering workflow described above provides three categorical variables (MM*_55*, *MM_65*, MM_70), each of which denotes the multimorbidity cluster a participant is assigned to in each age range (e.g. a participant could belong to a cardiometabolic cluster in the 0-55 age range, and then to a mental health cluster in the 55-65 age range depending on the new conditions they acquired from age 55 to 65). We included incident dementia as the outcome variable and multimorbidity cluster as the predictor of interest, and covaried for age at baseline, sex, and years of education. We first performed this analysis for each age range separately, irrespective of participants’ multimorbidity patterns at other age ranges (i.e. three separate models, each containing only one of MM_55, MM_65, MM_70). Then, to assess the impact of multimorbidity over the lifespan, we performed a logistic regression with incident dementia as outcome, all three of MM_55, MM_65, and MM_70 as predictors, and age, sex, and education as covariates. For this analysis, individuals excluded in the 65-70 years clustering due to unavailability of health records were all assigned to a separate group (EXCLUDE) at age 70. Throughout each analysis, a group representing low multimorbidity load (i.e. a relatively healthy group) was used as the reference group for statistical comparisons. P values were corrected for multiple comparisons using false discovery rate correction (𝑃𝐹𝐷𝑅 < 0.05 considered significant).

### Data and Code Availability

The data used in this study are available from the UK Biobank (https://www.ukbiobank.ac.uk/enable-your-research/apply-for-access). As restrictions apply to the availability of these data, which were used under licence for the current study, the authors cannot publicly share this data. The use of data from the UK Biobank was approved by the UK Biobank Access Committee (Project No. 95758). Code for this project is available at https://git.fmrib.ox.ac.uk/psp365/mm_risk.

